# Increased Risk of Poor Clinical Outcome in COVID-19 Patients with Diabetes Mellitus and in-hospital Mortality Predictors: A Retrospective Cohort from a Tertiary Hospital in Indonesia

**DOI:** 10.1101/2021.12.30.21266217

**Authors:** Md Ikhsan Mokoagow, Dante Saksono Harbuwono, Ida Ayu Kshanti, C Martin Rumende, Imam Subekti, Kuntjoro Harimurti, Khie Chen Lie, Hamzah Shatri

## Abstract

**Aim:** To determine association between diabetes in confirmed cases of COVID-19 and intensive care admission and in-hospital mortality, evaluate several laboratory parameters as mortality predictor, and develop predictors of in-hospital mortality among diabetics with COVID-19.

**Methods:** This retrospective cohort recruited all cases of COVID-19 hospitalized in Fatmawati General Hospital during March to October 2020. Inclusion criteria was RT-PCR confirmed cases of COVID-19 who aged 18 years and older while exclusion criteria were incomplete medical record or cannot be found and pregnant women.

**Results:** We enrolled 506 participants to this study with median age of 51 years (IQR:22), female (56.32%), and diabetes (28.46%). Diabetes increased intensive care admission (adjusted OR:6.07;95%CI:3.52-10,43) and in-hospital mortality (adjusted OR:50;95%CI:1.61-3.89). In predicting in-hospital mortality, ferritin and lactate dehydrogenase offered an acceptable discrimination, AUC:0.71 (95%CI: 0.62-0.79) and AUC:0.70 (95%CI: 0.61-0.78), respectively. The optimal cut-off of predicting mortality for ferritin was 786 g/mL and for LDH was 514.94 u/L. Factors include age above 70 years old, RBGs level on admission above 250 mg/dL or below 140 mg/dL, ferritin level above 786 ng/mL, and presence of ARDS increased the odds of mortality among individuals with diabetes.

**Conclusions:** Diabetes increases risk of intensive care admission and in hospital mortality in COVID-19. Multivariate analysis showed that older age, RBG on admission, high ferritin level, presence of ARDS increased the odds of mortality among individuals with diabetes.

## INTRODUCTION

Coronavirus Disease-2019 (COVID-19) first identified in Wuhan China and subsequently spread worldwide hence emerged as global health problem. Currently its case fatality rate in Indonesia exceeded the global rate.^1^ Presence of comorbidity contribute to poor clinical outcome.^2^ COVID-19 patients with diabetes are at risk of developing poor outcome during hospitalization.^3,4^ Our preliminary data in early pandemic documented half of individuals with diabetes admitted to our hospital due to COVID-19 were in critical diseases with a mortality rate of over 60% in this group.^5^ Identifying individuals with diabetes who are at higher risk of having clinical deterioration is of clinical importance.

Laboratory findings as predictors for poor outcome in COVID-19 has been extensively studied. Parameters such as C-reactive protein, lactate dehydrogenase, ferritin, D-dimer, neutrophils to lymphocyte ratio and monocyte to lymphocyte ratio have been associated with poor clinical outcome in COVID-19.^6-17^

It is conceivable that diabetes may contribute to poor clinical outcome and laboratory findings may predict in hospital mortality. Nevertheless, studies relating these occurrence remains limited. We sought to determine the association between diabetes in confirmed cases of COVID-19 and intensive care admission and in-hospital mortality, evaluate several laboratory parameters as mortality predictor, and develop predictors of in-hospital mortality among diabetics with COVID-19.

## 2. METHODS

### 2.1. Study design and participant

This is a single-centre, retrospective cohort study conducted in Fatmawati General Hospital from March to October 2020. Fatmawati General Hospital is a tertiary hospital located in South Jakarta. This hospital served as a referral hospital for severe COVID-19 for three consecutive cities, including South Jakarta, Depok, and South Tangerang cities (total population of 6,343,000). A total of 1,872 COVID-19 patients (confirmed and probable) were admitted during the study period. We included all COVID-19 confirmed individuals diagnosed based on RT-PCR test of nasal or oral-pharyngeal swab specimen, aged above 18 years or older, and at least had one measurement of random blood glucose test during admission. Individuals who were pregnant and data for the outcome cannot be achieved excluded from this study.

### 2.2 Outcomes

We reviewed demographical data, comorbidities, and laboratory findings based on patients’ medical charts. Diabetes was determined based on the patient’s medical history and laboratory parameters, including HbA1C ≥ 6.5 or fasting blood glucose ≥ 126 mg/dL or random blood glucose ≥ 200 mg/dL in two consecutive measurements. Comorbidity was determined based on the patient’s medical history and categorized into without comorbidity, present of 1 comorbidity, and present two or more comorbidity. Laboratory findings consisted of random blood glucose test (RBGs), C-reactive protein (CRP), Ferritin, D-dimer, lactate dehydrogenase (LDH), Neutrophile to lymphocyte ratio (NLR), and monocyte to lymphocyte ratio (MLR), all obtained at the time of admission. The disease severity was categorized based on WHO COVID-19 severity.1

Acute respiratory distress syndrome (ARDS) was determined based on oxygen saturation measured by pulse oximetry/ FiO2 ratio < 315 at the time of admission. We assessed the incidence of intensive care treatment, and mortality of COVID-19 patients with or without prior diabetes condition treated in our hospital.

### 2.3 Statistical analysis

Numeric data were presented as a median and inter-quartile range (IQR) or mean and standard deviation (SD) as determined by the Shapiro-Wilk test. Categorical data were presented as frequency and percent. Laboratory data cut-off was obtained based on previously published research for predicting critical conditions in COVID-19 patients. The cut-off for laboratory findings were RBG 180 mg/dL, CRP 5.213 mg/dL, Ferritin 304.3 ng/mL, d-dimer 2,010 ng/mL, LDH 353.5 u/L, NLR 8.085, MLR 0.364 respectively for intensive care outcome, and RBG 180 mg/dL, CRP 2.587 mg/dL, Ferritin 163.5 ng/mL, d-dimer 565 ng/mL, LDH 277 u/L, NLR 5.87, MLR 0.30 respectively for mortality outcome. Bivariate analysis using the chi-square test and multivariate logistics regression was performed to obtain crude and adjusted odds ratios. We assessed the association between diabetes and outcome using causative models. Nearly all study variables were assessed for their relationship with the outcomes. Backward selection for selecting covariates as confounding factors was used. Suppose the exclusion of covariate changes the OR crude of diabetes by more than 10% than the covariates is considered a potential confounder. If there is a clinically meaningful relationship, the potential confounders are regarded as confounders, and adjusted ORs of association between diabetes and outcomes were obtained. We measured the performance of laboratory findings by measuring the area under curve, sensitivity, and specificity. The optimal cut-off value is measured by determining the closest distance between the point (0,1) and the point on the ROC curve (d) and determining the farthest vertical distance between the line of equality and the point on the ROC curve (Youden index). All the analysis was performed using STATA Version 12. The study was reviewed and approved by the Research Ethic Committee of Fatmawati General Hospital (27/KEP/XII/2020).

## 3. RESULTS

### 3.1 Patients characteristics

During the study period from 11 March to 31 November 2020, a total of 616 confirmed COVID-19 patients were admitted to our hospital. Of all of them, only 600 complete medical records can be obtained. We excluded children (27 persons), pregnant women (65 persons), and those who never had blood glucose measurements during admission (6 persons). Therefore, a total of 506 participants were included in this study (Figure 1)

**Figure 1.**
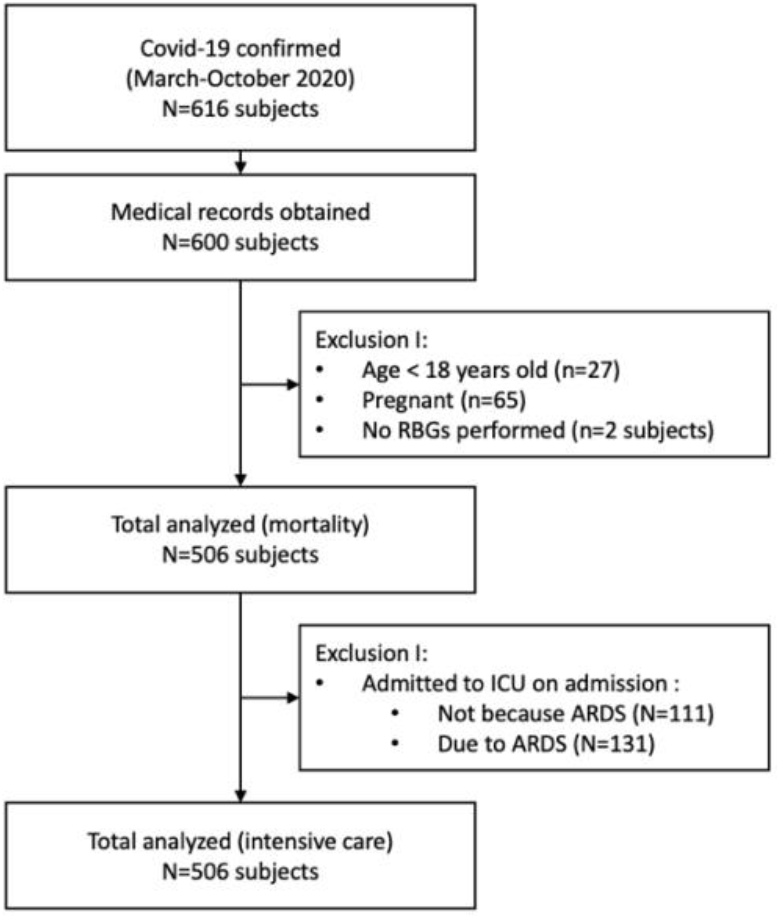
Flow Diagram

The characteristic of the participants is presented in Table 1. Diabetes was found in 28.46% of individuals. Most of the subjects in this study were below 60 years old, with a median (IQR) age 51 (22) years old. Females are more prevalent compare to males (56.32 vs. 43.68%). Despite diabetes, the most common comorbidity found in this study was hypertension (30.80%) and chronic kidney disease (24.20%).). The proportion of severity levels among subjects at the admission time was as follows; mild 11.07%, moderate 34.39%, severe 8.89%, and critical 45.85% (ARDS 44.66%, sepsis 1.19%, septic shock 6.72%). The mean hospital duration in this study was 13 (12 days).

**Table 1.** Characteristics of subjects

| Variable | Total | Diabetes |  |
| --- | --- | --- | --- |
|  |  | Yes<br>144 (28,46) | No<br>362 (71,54) |
| Age, years old – Median (IQR) | 51 (22) | 57 (15) | 48,5 (22) |
| Age Category |  |  |  |
| ≥ 60 years old | 143 (28,26) | 60 (41,67) | 83 (22,93) |
| < 60 years old | 363 (71,47) | 84 (58,33) | 279 (77,07) |
| Sex |  |  |  |
| Male – n (%) | 221 (43,68) | 67 (46,53) | 154 (42,54) |
| Female – n (%) | 285 (56,32) | 77 (53,47) | 208 (57,46) |
| Comorbidity |  |  |  |
| Hypertension – n (%) | 154 (30,80) | 60 (42,25) | 94 (26,6) |
| Heart disease – n (%) | 57 (11,40) | 22 (15,49) | 35 (9,78) |
| Chronic obstructive pulmonary disease – n (%) | 23 (4,60) | 6 (4,23) | 17 (4,75) |
| Stroke – n (%) | 21 (4,20) | 5 (3,52) | 16 (4,47) |
| Chronic kidney disease – n (%) | 121 (24,20) | 43 (30,28) | 78 (21,79) |
| Liver Disease – n (%) | 5 (0,99) | 1 (0,69) | 4 (1,10) |
| Other immunodeficiency – n (%) | 19 (3,80) | 8 (5,64) | 11 (3,08) |
| Laboratory parameters |  |  |  |
| RBGs, mg/dL – median (IQR) | 109 (54) | 139,5 (122) | 105 (41) |
| CRP, mg/dL – median (IQR) | 4,5 (10,9) | 6,5 (13,5) | 3,97 (9,2) |
| Feritin, ng/mL – median (IQR) | 542,5 (1172) | 772,5 (1235,52) | 474 (1043) |
| D-dimer, ng/mL – median (IQR) | 1570 (2260) | 2260 (3315,8) | 1390 (2390,42) |
| LDH, u/L – median (IQR) | 485,49 (322) | 523,5 (378,5) | 477,14 (306,99) |
| NLR – median (IQR) | 6,42 (5,41) | 6,38 (5,45) | 6,51 (5,45) |
| MLR – median (IQR) | 0,49 (0,34) | 0,45 (0,272) | 0,5 (0,363) |
| Hospital duration, day – median (IQR) | 13 (12) | 15 (16) | 13 (8) |

### 3.2 The association between diabetes and outcome

Of all of the subjects included, 297 (58.70%) subjects were admitted to the intensive care unit, while 140 (27.67%) subjects were dead during admission. Among subjects with diabetes versus subjects without diabetes, a higher proportion was admitted to intensive care (86.11% vs. 47.79%, respectively p<0.001, supplementary 1) and death (45.14% vs. 20.72%, respectively p<0.001, supplementary 2).

In multivariate logistic regression analysis, diabetes was associated with increased odds of admitted on intensive care OR 6.07 (95% CI: 3.52-10.43) (fig.2), and death OR 2.50 (95% CI: 1.61-3.89) (fig.3) after adjustment with age, sex, comorbidity, random blood glucose levels, C-reactive protein, ferritin, d-dimer, lactate dehydrogenase levels, neutrophile and monocyte to lymphocyte ratio levels.

**Figure 2.**
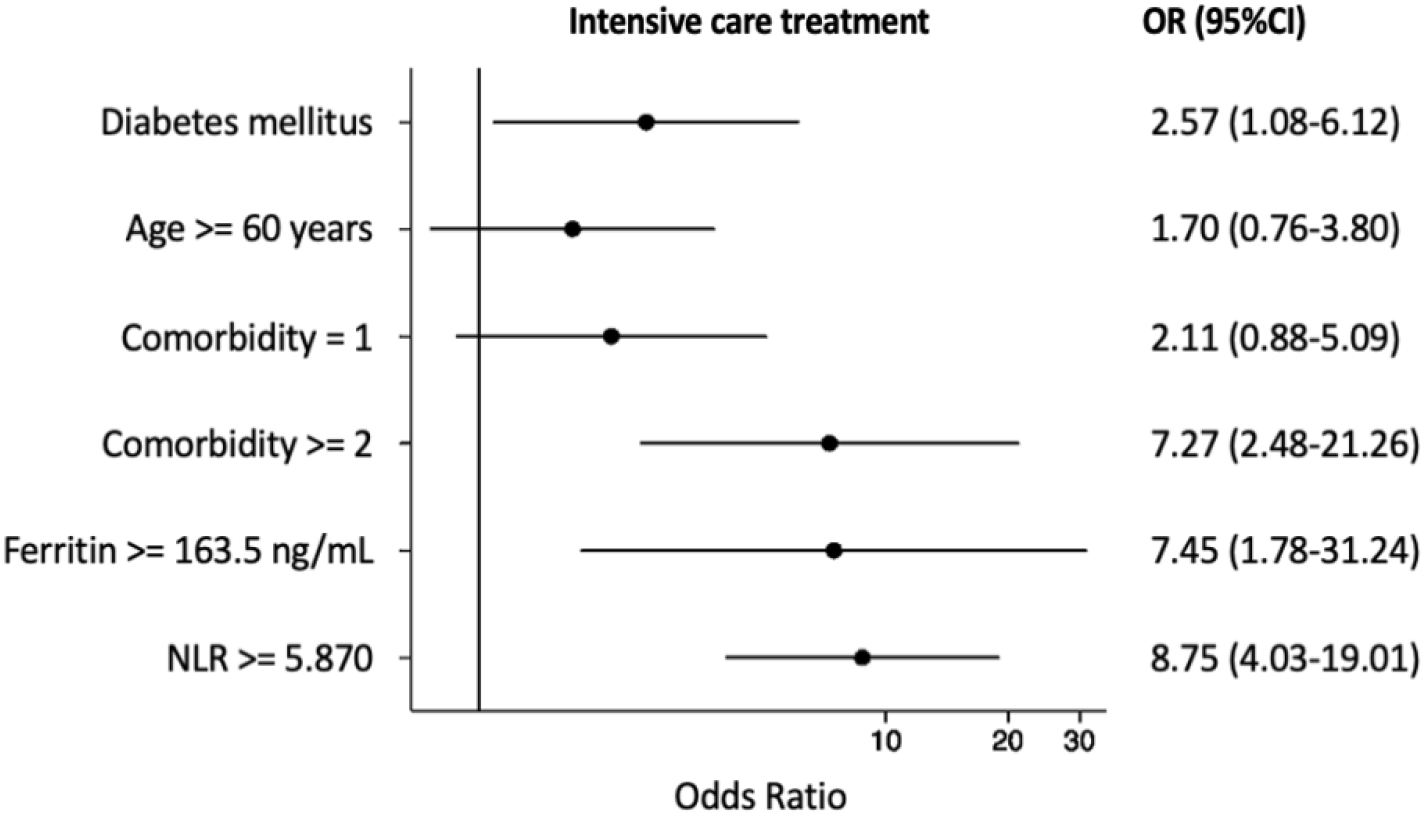
The Association between Diabetes and Intensive care treatment in COVID-19 patients

**Figure 3.**
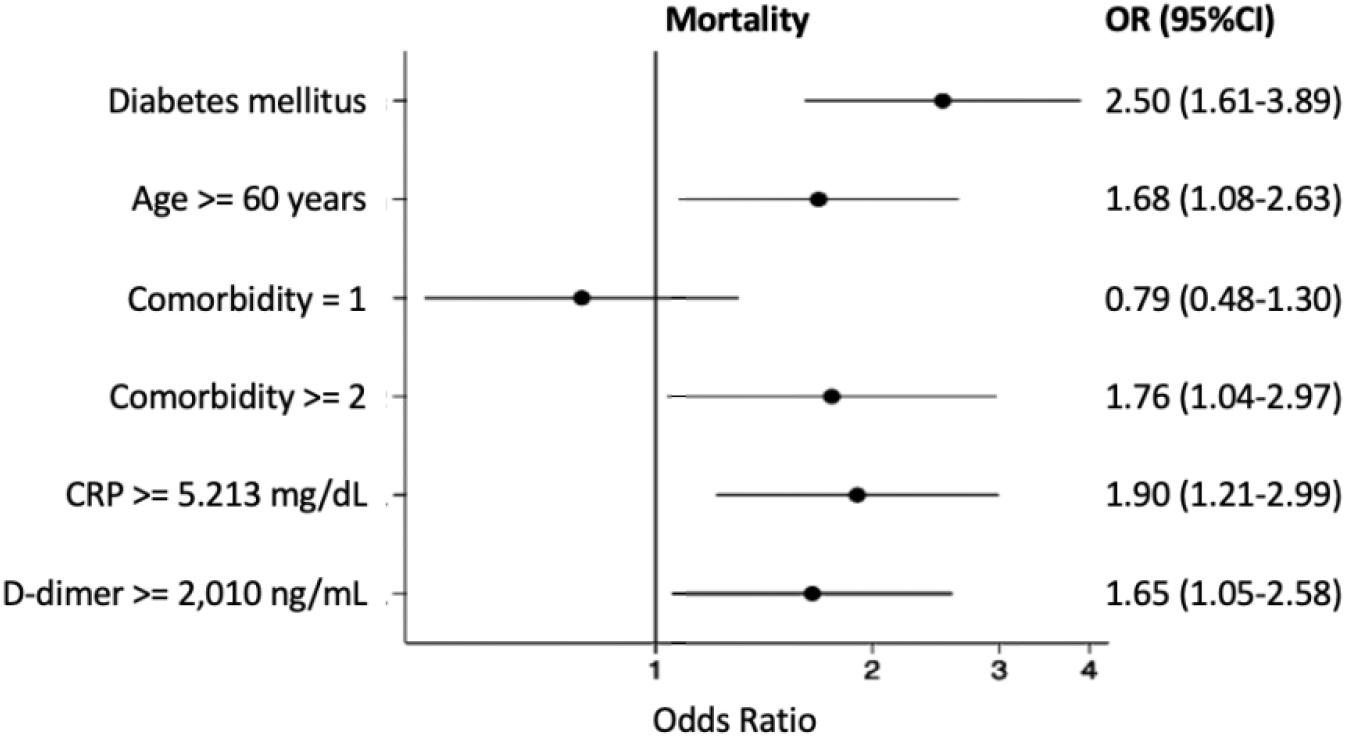
The Association between Diabetes and Mortality in COVID-19 patients

### 3.3. Laboratory parameters as a mortality predictor in diabetes subjects

The accuracy of laboratory parameters to predict mortality in diabetes subjects was shown based on the area under the curve (AUC) of each parameter. All of the laboratory parameters had AUC below 0.70 in predicting mortality of diabetes subjects except for ferritin (AUC: 0.71, 95%CI: 0.62-0.79) and LDH (AUC: 0.70, 95%CI: 0.61-0.78). The optimal cut-off for ferritin to predict mortality among subjects with diabetes was 786 ng/mL (sensitivity: 0.72, specificity: 0.68) and the optimal cut off for LDH was 514.94 u/L (sensitivity: 0.71, specificity: 0.62) (Table 2).

**Table 2.** Laboratory parameters as a mortality predictor in COVID-19 with diabetes subjects

| Laboratory parameter | AUC | Cut-off* | Spec* | Sens | Spec | PPV | NPV | LR+ | LR- |
| --- | --- | --- | --- | --- | --- | --- | --- | --- | --- |
| CRP (mg/dL) | 0.650 | ≥ 5,213 | 0,710 | 0,621 | 0,601 | 0,373 | 0,806 | 1,556 | 0,631 |
| LDH (u/L) | 0.675 | ≥ 353,5 | 0,890 | 0,871 | 0,262 | 0,311 | 0,842 | 1,180 | 0,492 |
| Ferritin (ng/mL) | 0.662 | ≥ 304,3 | 0,706 | 0,779 | 0,352 | 0,315 | 0,806 | 1,202 | 0,628 |
| d-dimer (ng/mL) | 0.645 | ≥ 2.010 | 0,740 | 0,564 | 0,637 | 0,373 | 0,793 | 1,554 | 0,684 |
| **NLR** | 0.500 | $\geq 8,085$ | 0,848 | 0,221 | 0,762 | 0,263 | 0,719 | 0,929 | 1,022 |
| **MLR** | 0.503 | $\geq 0,364$ | 0,800 | 0,643 | 0,383 | 0,287 | 0,740 | 1,042 | 0,932 |
| **RBGs (mg/dL)** | 0.557 | $\geq 180$ | - | 0,271 | 0,861 | 0,427 | 0,755 | 1,950 | 0,847 |

### 3.4 Factors associated with mortality among diabetes subjects

We explore factors affecting mortality in diabetes subjects. Based on bivariate analysis, some variables have a statistically significant association with mortality. They were random blood glucose levels, CRP, Ferritin, LDH, D-dimer, and ARDS (supplementary table 4). The multivariate analysis was performed. Based on multivariate analysis, it is seen that age above 70 years old, RBGs level above 250 mg/dL or below 140 mg/dL, ferritin level above 786 ng/mL, and presence of ARDS increased the odds of mortality among subjects with diabetes. Meanwhile, comorbidity, NLR above 4.69, and MLR above 0.8 are confounding factors that attenuate the other risks (Table 3).

**Table 3.** Risk Factor of mortality among COVID-19 with diabetes subjects.

| Variable | Coefficient B | p-value | OR | 95%CI |
| --- | --- | --- | --- | --- |
| **RBGs mg/dL** |  |  |  |  |
| < 140 | 1,618 | 0,003 | 5,04 | 1,72-14,78 |
| > 250 | 1,768 | 0,006 | 5,86 | 1,65-20,78 |
| **Age $\geq 70$ years old** | 0,283 | 0,033 | 1,33 | 1,02-1,72 |
| **NLR $\geq 4.69$** | 0,660 | 0,130 | 1,94 | 0,82-4,55 |
| **Ferritin $\geq 786$ ng/mL** | 1,731 | 0,000 | 5,65 | 2,40-13,29 |
| **ARDS** | 1,198 | 0,009 | 3,31 | 1,35-8,11 |
| **Constanta** | -3,498 | 0,000 | 0,04 | 0,01-0,12 |

## 4. DISCUSSION

Our study is the first study in Indonesia that clinically analyzed the relationship between DM and COVID-19 outcomes. We found that the proportion of DM among COVID-19 confirmed patients admitted to our hospital was 28.5%. Furthermore, DM increased the risk of having intensive care treatment by 6.07, and death by 2.50 compared to those who did not have DM. This finding is consistence with other findings including a meta-analysis who included 76 studies, in which 10 studies (for severe outcome) and 8 studies (for mortality outcome) came from outside China. The risk of having severe condition increased by 2.02 (95 CI: 1.44-2.84, I^2^: 55%, P<0.02) and the risk of having mortality increased by 2.21 (95 CI: 1.83-2.66, I^2^: 50%, P<0.01) in DM patients compared to those who did not had DM outside China.^18^ Other meta-analysis including in hospital adults’ patients with COVID-19 found the same. The risk of in hospital mortality increased by 2.39 (95 CI: 1.65-3.46, I^2^: 62%, P:0.001).^19^ However, those meta-analysis failed to prove diabetes increased the risk of severe conditions.^19^ Other meta-analysis including three study analyzed intensive care treatment among COVID-19 also failed to prove diabetes increased the risk of having intensive care treatment. (OR: 1.47, 95 CI: 0.38-5.67, I^2^: 63%, P: 0.07).^20^

The results of previous meta-analysis studies are still conflicting and had high heterogeneity. In this study, the OR of the relationship between diabetes and intensive care was high, 6.07. However, if we looked at the confidence interval in this relationship (3.52-10.43), the confidence interval is wide. Some reasons can explain this finding. First, besides clinical consideration, physicians’ decision to admit subjects to an intensive care facility was affected by beds availability and family consent. Also, a small sample size might give a wide range of confidence intervals. From all 606 subjects admitted to our hospital, we could assess only 264 subjects (42.86%) after the exclusion. Lastly, we used OR rather than Risk Ratio (RR) as an association measurement because we used logistic regression for multivariate analysis. Therefore, we can assume that this relationship is not strong and is most likely affected by chance. Further research with clinically objective outcomes like ARDS, sepsis, or other critical conditions is recommended.

Besides of COVID-19, it is well known that subjects with diabetes are prone to have infections compared to non-diabetes subjects. There is also paucity evidence in epidemiological studies where DM patients had increased risk of having mortality by COVID-19.^18-20^ The potential mechanisms that could explain the high COVID-19 mortalities among DM subjects were impaired immune responses, glycemic instability, chronic subclinical inflammation, and the presence of associated comorbidities in diabetes subjects.^21^

Several abnormal immune system have been seen in diabetes subjects, those including higher affinity cellular binding for virus entry, inhibition of viral clearance, impaired T-cell function, lymphopenia, and exaggerated inflammatory response associated with an increased renin-angiotensin system (RAS) activation in several tissues.^22,23^ In addition, those conditions were worsened by chronic glycemic instability and association inflammation, which contribute to increased susceptibility to hyperinflammation and cytokine storm.^23^ This findings also supported our study in which high ferritin (> 786 ng/mL) increased the risk of mortality.

To date, there is no available report regarding cut-off for laboratory findings in predicting mortality of COVID-19 patients with diabetes. In this study, among all the laboratory parameters, only ferritin shows good performance in predicting mortality. Ferritin gives an AUC score of 0.71 with an optimal cut-off at 786 ng/mL, giving it a sensitivity of 0.72 and specificity of 0.68. LDH seemed like a good predictor for mortality among subjects with diabetes. However, it failed to prove its association with mortality in multivariate analysis (supplementary table 4).

Many studies have been reported that ferritin is an independent factor of poor outcomes in COVID-19 patients.^24^ Ferritin is an intracellular protein that can store iron and plays a critical role in inflammatory diseases. It is an acute-phase protein, which is elevated during inflammation or infection. The condition of “hyperferritinemia syndrome” is proved to be associated with highly active diseases, resulting in immune activation and coagulation disturbances.^24^ Among COVID-19 patients, subjects with diabetes conditions already had a higher baseline of ferritin.^25,26^ The out-of-control inflammatory state in diabetes subjects who are already on impaired immune response may cause widespread tissue damage and susceptibility to poor outcomes.

Subjects with diabetes also have a greater risk for poor glycemic control. Acute diabetes complications were likely to occur (Diabetic ketoacidosis, Hyperglycemia hyperosmolar syndrome, and hypoglycemia), which then inhibits the ability to mitigate sepsis and higher mortality rate due to the complication itself.^23^ Furthermore, the presence of associated morbidities and the use of agents that can modulate angiotensin-converting enzyme 2 (ACE2) expression have been put forth to explain the latter association.^22^ We demonstrated that age and random blood glucose levels on admission were the independent risk factors of mortality in COVID-19 subjects with diabetes. We found age is major role when diabetes subjects reached 70 years or older. Also, subjects with RBGs levels above 250 mg/dL or below 140 mg/dL were found at increased risk of death.

Following our findings, a meta-analysis including 22 studies showed that individuals with a more severe diabetes condition have a poorer prognosis of COVID-19 compared to those with milder diabetes conditions. ^27^ A Spanish COVID-19 registry found admission hyperglycemia (RBG > 180 mg/dL) regardless of diabetes diagnosis was a strong predictor of all-cause mortality in non-critically hospitalized patients.^28^ Moreover, the chronic hyperglycemia condition also leads to increased fibrinogen amyloid changes, leading to hypercoagulability conditions.^29^ It is not surprising that hyperglycemia increased the risk of poor outcomes. Current evidence showed tight glucose control with insulin infusion in ICU had a lower risk of severe symptoms and mortality. It is also well known that hypoglycemia might produce the same effect as acute hyperglycemia. Nevertheless, glucose variability also induced cytokines release and worsened prognosis. Currently, the optimal cut-off for glycemic control among COVID-19 patients with hyperglycemia remains elusive. It is challenging to maintain normal glycemic in COVID-19 patients with glycemia, and it was even more complicated with the use of corticosteroid treatment. Therefore, increased attention is needed for blood glucose regulations in COVID-19 patients. Further research regarding optimal glucose target and evidence of treatment is needed.

This study has several limitations to be discussed. First, we cannot analyze the time relationship in this study. That is because we did not measure the diabetes duration. Second, diabetes is a heterogeneous population. We included all subjects with diabetes conditions, including type 1 diabetes, type 2 diabetes, newly diagnosed diabetes, controlled or uncontrolled diabetes, those who received insulin or oral diabetes medication. Therefore, there is no specificity in this study, nor we performed a sub-analysis of which of these different factors might play a role in the outcome. Lastly, our study is conducted in a tertiary facility and one of the largest hospitals in Indonesia that the government-appointed as an advanced referral hospital. About 45.85% of subjects in this study already came with critical conditions, and 8.89% were already in severe conditions. Therefore, readers should be careful in generalizing our results.

In accordance with our findings, a meta-analysis including 22 studies showed individuals with a more severe diabetes condition have a poorer prognosis of COVID-19 compared to those with milder diabetes conditions.^27^ A Spanish COVID-19 registry found admission hyperglycemia (RBG > 180 mg/dL) regardless of diabetes diagnosis was a strong predictor of all-cause mortality in non-critically hospitalized patients.^28^ Moreover, the chronic hyperglycemia condition also leads to increased fibrinogen amyloid changes, leading to hypercoagulability conditions.^29^ It is not surprising that hyperglycemia increased the risk of poor outcome.^30^ Also, hypoglycemia might produce the same effect as acute hyperglycemia. The optimal cut-off for COVID-19 patients with hyperglycemia treatment remains elusive. Current evidence showed tight glucose control with insulin infusion in ICU had a lower risk of severe symptoms and mortality.^31^ But glucose variability also induced cytokines release and worsen prognosis.^31^ It is challenging to maintain normal glycemic in COVID-19 patients with glycemia and it was even harder with the use of corticosteroid treatment. Increased attention is needed for blood glucose regulations in COVID-19 patients. Further research regarding optimal glucose target and evidence of treatment is needed.

This study has several limitations to be discussed. First, we cannot analyze the time relationship in this study. That is because we did not measure the diabetes duration. Second, basically diabetes is heterogenous population, we included all subjects with diabetes conditions, including type 1 diabetes, type 2 diabetes, newly diagnosed diabetes, controlled or uncontrolled diabetes, those who received insulin or oral diabetes medication. Therefore, there is no specificity in this study, nor we performed a sub-analysis which of these different factors might play role in the outcome. Lastly, our study is conducted in a tertiary facility and one of the largest hospitals in Indonesia that the government-appointed as an advanced referral hospital. About 45.85% of subjects in this study already came with critical conditions, and 8.89% were already in severe conditions. Therefore, readers should be careful in generalizing our results.

## CONCLUSION

The increased risk of intensive care and mortality in people with diabetes mellitus who experience COVID-19 requires special attention from various stakeholders, including clinicians, hospitals, the government, and the community. In addition, suitable clinical and laboratory monitoring protocols are needed to anticipate early clinical deterioration.

## Data Availability

All data produced in the present study are available upon reasonable request to the authors.

## ACKNOWLEDGEMENTS

The authors would like to acknowledge COVID-19 Management Tasks Force, Fatmawati General Hospital for their tremendous endeavour during the pandemic.

---

**Supp 1.**
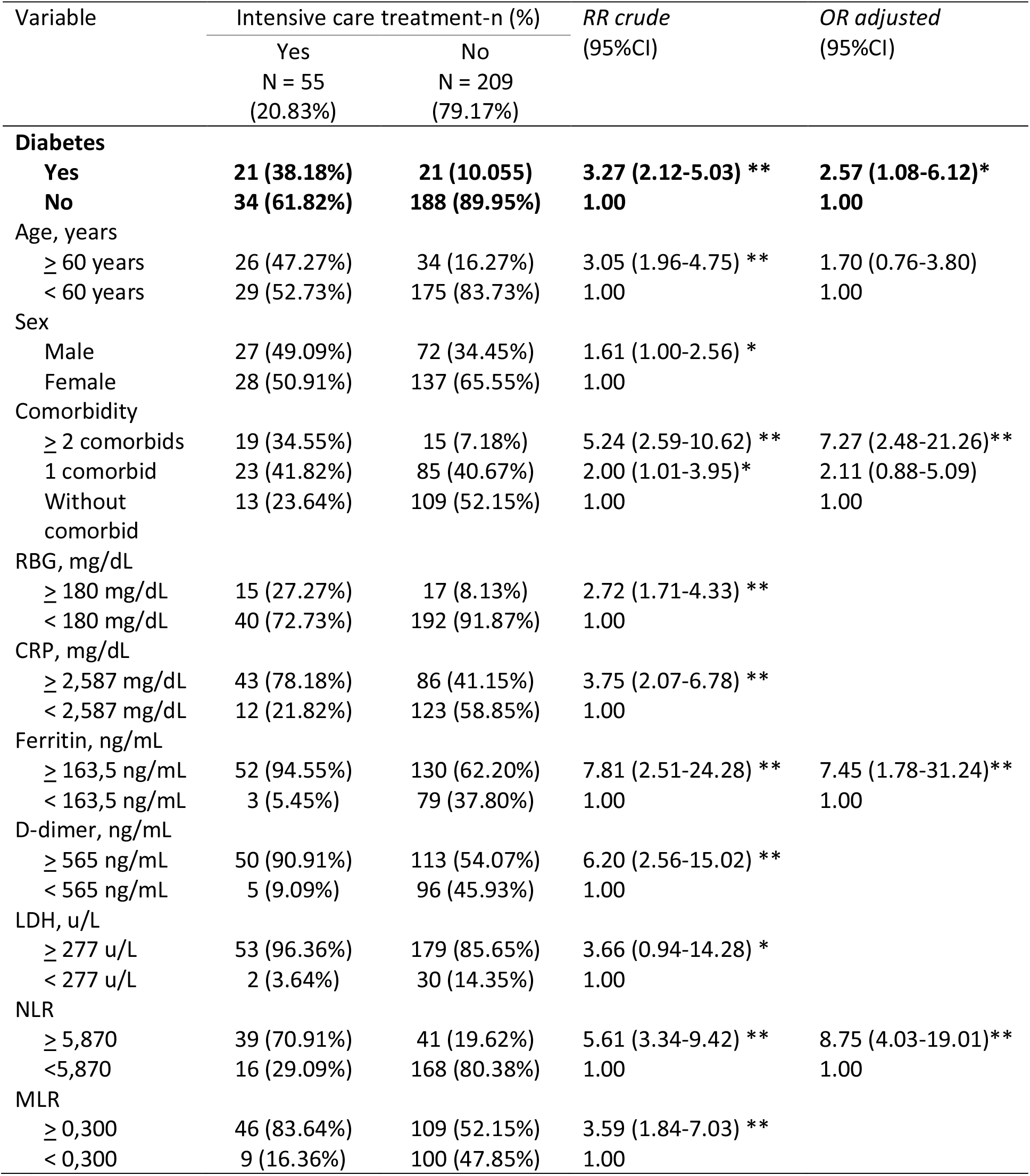
The Association between Diabetes and intensive care treatment in COVID-19 patients

| Variable | Intensive care treatment-n (%) |  | *RR crude*<br>(95%CI) | *OR adjusted*<br>(95%CI) |
| --- | --- | --- | --- | --- |
|  | Yes<br>N = 55<br>(20.83%) | No<br>N = 209<br>(79.17%) |  |  |
| **Diabetes** |  |  |  |  |
| **Yes** | **21 (38.18%)** | **21 (10.055)** | **3.27 (2.12-5.03) \*\*** | **2.57 (1.08-6.12)\*** |
| **No** | **34 (61.82%)** | **188 (89.95%)** | **1.00** | **1.00** |
| Age, years |  |  |  |  |
| ≥ 60 years | 26 (47.27%) | 34 (16.27%) | 3.05 (1.96-4.75) ** | 1.70 (0.76-3.80) |
| < 60 years | 29 (52.73%) | 175 (83.73%) | 1.00 | 1.00 |
| Sex |  |  |  |  |
| Male | 27 (49.09%) | 72 (34.45%) | 1.61 (1.00-2.56) * |  |
| Female | 28 (50.91%) | 137 (65.55%) | 1.00 |  |
| Comorbidity |  |  |  |  |
| ≥ 2 comorbids | 19 (34.55%) | 15 (7.18%) | 5.24 (2.59-10.62) ** | 7.27 (2.48-21.26)** |
| 1 comorbid | 23 (41.82%) | 85 (40.67%) | 2.00 (1.01-3.95)* | 2.11 (0.88-5.09) |
| Without comorbid | 13 (23.64%) | 109 (52.15%) | 1.00 | 1.00 |
| RBG, mg/dL |  |  |  |  |
| ≥ 180 mg/dL | 15 (27.27%) | 17 (8.13%) | 2.72 (1.71-4.33) ** |  |
| < 180 mg/dL | 40 (72.73%) | 192 (91.87%) | 1.00 |  |
| CRP, mg/dL |  |  |  |  |
| ≥ 2,587 mg/dL | 43 (78.18%) | 86 (41.15%) | 3.75 (2.07-6.78) ** |  |
| < 2,587 mg/dL | 12 (21.82%) | 123 (58.85%) | 1.00 |  |
| Ferritin, ng/mL |  |  |  |  |
| ≥ 163,5 ng/mL | 52 (94.55%) | 130 (62.20%) | 7.81 (2.51-24.28) ** | 7.45 (1.78-31.24)** |
| < 163,5 ng/mL | 3 (5.45%) | 79 (37.80%) | 1.00 | 1.00 |
| D-dimer, ng/mL |  |  |  |  |
| ≥ 565 ng/mL | 50 (90.91%) | 113 (54.07%) | 6.20 (2.56-15.02) ** |  |
| < 565 ng/mL | 5 (9.09%) | 96 (45.93%) | 1.00 |  |
| LDH, u/L |  |  |  |  |
| ≥ 277 u/L | 53 (96.36%) | 179 (85.65%) | 3.66 (0.94-14.28) * |  |
| < 277 u/L | 2 (3.64%) | 30 (14.35%) | 1.00 |  |
| NLR |  |  |  |  |
| ≥ 5,870 | 39 (70.91%) | 41 (19.62%) | 5.61 (3.34-9.42) ** | 8.75 (4.03-19.01)** |
| <5,870 | 16 (29.09%) | 168 (80.38%) | 1.00 | 1.00 |
| MLR |  |  |  |  |
| ≥ 0,300 | 46 (83.64%) | 109 (52.15%) | 3.59 (1.84-7.03) ** |  |
| < 0,300 | 9 (16.36%) | 100 (47.85%) | 1.00 |  |

**Supp 2.** The Association between Diabetes and mortality in COVID-19 patients

| Variable | Mortality- n(%) |  | *RR crude*<br>(95%CI) | *OR adjusted*<br>(95%CI) |
| --- | --- | --- | --- | --- |
|  | Yes<br>N = 140<br>(27.67%) | No<br>N = 366<br>(72.33%) |  |  |
| **Diabetes** |  |  |  |  |
| **Yes** | **65 (45,14)** | **79 (54,86)** | **2.18 (1.66-2.86) \*\*** | **2,50 (1.61-3,89) \*\*** |
| **No** | **75 (20,72)** | **287 (79,28)** | **1,00** | **1,00** |
| Age, years |  |  |  |  |
| ≥ 60 years | 54 (37,76) | 89 (62,24) | 1.59 (1.21-2.19) ** | 1,68 (1,08-2,63) * |
| < 60 years | 86 (23,69) | 277 (76,31) | 1,00 | 1,00 |
| Sex |  |  |  |  |
| Male | 71 (32,13) | 150 (67,87) | 1,32 (0.99-1.74) |  |
| Female | 69 (24,21) | 216 (75,79) | 1,00 |  |
| Comorbidity |  |  |  |  |
| ≥ 2 comorbid | 44 (42,31) | 60 (57,69) | 3.23 (2.03-5.14) ** | 1,76 (1,04-2,97) * |
| 1 comorbid | 41 (24,70) | 125 (75,30) | 1.95 (1.23-3.10) | 0,79 (0,48-1,30) |
| Without comorbid | 55 (23,31) | 181 (76,69) | 1,00 | 1,00 |
| RBG, mg/dL |  |  |  |  |
| ≥ 180 mg/dL | 38 (42,70) | 51 (57,30) | 1.96 (1.48-2.59) ** |  |
| < 180 mg/dL | 102 (24,46) | 315 (75,54) | 1,00 |  |
| CRP, mg/dL |  |  |  |  |
| ≥ 5,213 mg/dL | 87 (37,34) | 146 (62,66) | 3.76 (2.61-5.40) ** | 1,90 (1,21-2,99) ** |
| < 5,213 mg/dL | 53 (19,41) | 220 (80,59) | 1,00 | 1,00 |
| Ferritin, ng/mL |  |  |  |  |
| ≥ 304,3 ng/mL | 109 (31,50) | 237 (68,50) | 1,91 (1,20-3,18) ** |  |
| < 304,3 ng/mL | 31 (19,38) | 129 (80,62) | 1,00 |  |
| d-dimer, ng/mL |  |  |  |  |
| ≥ 2.010 ng/mL | 79 (37,26) | 133 (62,74) | 3.89 (2.27-6,65) ** | 1,65 (1,05-2,58) * |
| < 2.010 ng/mL | 61 (20,75) | 233 (79,25) | 1,00 | 1,00 |
| LDH, u/L |  |  |  |  |
| ≥ 353,5 u/L | 122 (31,12) | 270 (68,88) | 3.06 (2.27-4.12) ** |  |
| < 353,5 u/L | 18 (15,79) | 96 (84,21) | 1,00 |  |
| NLR |  |  |  |  |
| ≥ 8,085 | 31 (26,27) | 87 (73,73) | 3,27 (2.46-4.36) ** |  |
| < 8,085 | 109 (28,09) | 279 (71,91) | 1,00 |  |
| MLR |  |  |  |  |
| ≥ 0,364 | 90 (28,66) | 224 (71,34) | 2,78 (1,93-3.99) ** |  |
| < 0,364 | 50 (26,04) | 142 (73,96) | 1,00 |  |

